# Individual Deviations from Normative Electroencephalographic Connectivity Predict Antidepressant Response

**DOI:** 10.1101/2023.05.24.23290434

**Authors:** Xiaoyu Tong, Hua Xie, Wei Wu, Corey Keller, Gregory Fonzo, Matthieu Chidharom, Nancy Carlisle, Amit Etkin, Yu Zhang

**Author notes:** Corresponding author: Yu Zhang Assistant Professor of Bioengineering Lehigh University Bethlehem, PA 18015, USA.

## Abstract

Antidepressant medications yield unsatisfactory treatment outcomes in patients with major depressive disorder (MDD) with modest advantages over the placebo. This modest efficacy is partly due to the elusive mechanisms of antidepressant responses and unexplained heterogeneity in patient’s response to treatment — the approved antidepressants only benefit a portion of patients, calling for personalized psychiatry based on individual-level prediction of treatment responses. Normative modeling, a framework that quantifies individual deviations in psychopathological dimensions, offers a promising avenue for the personalized treatment for psychiatric disorders. In this study, we built a normative model with resting-state electroencephalography (EEG) connectivity data from healthy controls of three independent cohorts. We characterized the individual deviation of MDD patients from the healthy norms, based on which we trained sparse predictive models for treatment responses of MDD patients. We successfully predicted treatment outcomes for patients receiving sertraline (r = 0.43, p < 0.001) and placebo (r = 0.33, p < 0.001). We also showed that the normative modeling framework successfully distinguished subclinical and diagnostic variabilities among subjects. From the predictive models, we identified key connectivity signatures in resting-state EEG for antidepressant treatment, suggesting differences in neural circuit involvement between treatment responses. Our findings and highly generalizable framework advance the neurobiological understanding in the potential pathways of antidepressant responses, enabling more targeted and effective MDD treatment.

**Trial Registration:** Establishing Moderators and Biosignatures of Antidepressant Response for Clinical Care for Depression (EMBARC), NCT#01407094

## Introduction

Major depressive disorder (MDD) is one of the most common psychiatric disorders and the leading cause of ill health and disability worldwide, with an over 20% lifetime prevalence (1). Despite the high incidence, neuropathological mechanisms underlying MDD are still in debate (2–5), hampering the development of antidepressants and yielding unsatisfactory treatment outcomes and low life quality of MDD patients. In fact, although numerous medications have been approved for MDD treatment, their effects over placebo are modest at best, with only a few newly approved medications showing equivocal efficacy (6–11). This unsatisfactory response rate is partly because MDD patients have varied responses to antidepressants due to underlying heterogeneity (12, 13). Additionally, an antidepressant may take at least 4 weeks to take effect and MDD patients may spend months to years searching through options before responding (14) while experiencing the side effects (15). Thus, knowing sooner whether a treatment will be effective for a particular patient would help psychiatrists move much more quickly through clinical decision trees. Therefore, one key step toward improved treatment outcome and life quality for MDD patients is to achieve precision medicine by employing pre-treatment measures to quantify the response-predictive individual deviations in psychopathological dimensions, as suggested by Research Domain Criteria (16) (RDoC).

In the journey towards precision medicine for psychiatric disorders, neuroimaging techniques have demonstrated their exceptional capabilities to identify neurobiological alterations predictive of disorder diagnosis (17–23). However, previous neuroimaging studies mainly focused on the group-level brain feature differences between patients and healthy controls, neglecting the heterogeneity within the patient group. To this end, a collection of studies have made efforts to identify subtypes within each disorder (24–28), aiming to achieve precision medicine by providing different treatments based on subtypes. However, these subtype findings have not yet been translated to improved clinical practice, partly because they still failed to construct a continuous spectrum of psychopathological dimensions to address the heterogeneity within subtypes, as well as the dissociation between the identified subtypes and treatment responses (29). To address these challenges, *normative modeling* is a promising framework to dissect patient heterogeneity and conceptualize psychopathological dimensions by quantifying individual deviations from a healthy norm (30). While a number of neuroimaging analyses have already employed the normative modeling framework, such as the development of continuous disease spectrums (31) and the quantification of heterogeneity within psychiatric disorders (32), they are mostly limited to the implementation using structural magnetic resonance imaging (MRI) (31, 32). Nevertheless, the underexplored functional connectivity (FC) features may be more advantageous as they capture the brain characteristics associated with general cognitive functioning that not necessarily lead to structural changes. Indeed, our prior work with resting-state functional MRI (rsfMRI) has shown the potential of FC-based normative modeling to quantify individual brain dysfunction and parse neurobiological heterogeneity in PTSD (33). Meanwhile, electroencephalography (EEG) provides another non-invasive neuroimaging technique, which in contrast to rsfMRI is cost-effective and easy-to-operate in clinical practice. Recent studies demonstrated that resting-state EEG (rsEEG) connectivity also facilitated MDD diagnosis (34) and revealed MDD subtypes (28). Taken together, these latest research efforts suggested the capability of FC-based individual deviations to provide essential information of the psychopathological dimensions, thus paving the way toward precision medicine for MDD.

In this study, we developed a rsEEG FC-based normative modeling framework to identify clinically translatable pre-treatment biomarkers for antidepressant responses. To this end, we constructed an EEG FC-based normative modeling framework that quantified individual deviations in psychopathological dimensions (Fig. 1) by leveraging three independent cohorts.

**Fig. 1.**
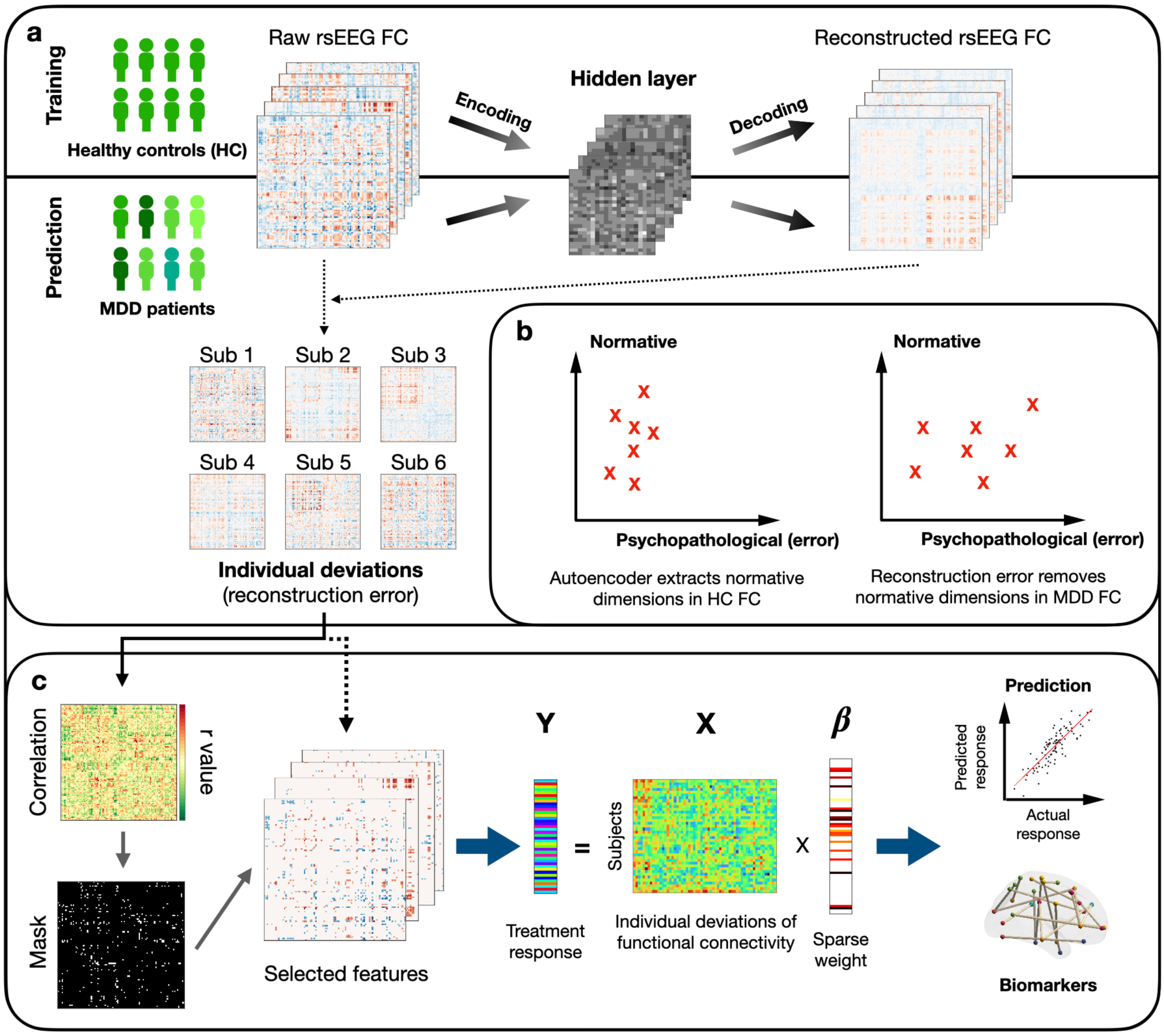
Flowchart of antidepressant response prediction based on individual deviations from normative electroencephalographic (EEG) functional connectivity (FC) **a** Normative modeling framework. First, a healthy norm was developed with healthy EEG FC using an autoencoder. Second, patient EEG FC were fed into the autoencoder, generating reconstructed EEG FC for each patient. The subject-level individual deviations from normative EEG FC were then quantified as the reconstruction error (raw FC - reconstructed FC) yielded by the autoencoder. **b** Distribution of individual deviations. The autoencoder captures common dimensions shared by healthy controls. The reconstruction errors, namely the individual deviations, reflect unseen dimensions in healthy controls. Those unseen dimensions were hypothesized to be associated with diseases, including MDD and other psychopathological dimensions. Thus, the individual deviations amplify the disease dimensions to yield FC features that are more predictive of treatment responses. **c** Predictive modeling framework. The individual deviations from normative EEG FC were utilized as the input features of the predictive modeling framework. First, correlation analyses were conducted on all individual deviations to identify statistically significant features of antidepressant responses. Second, insignificant features were masked out to reduce the feature dimensionality. Lastly, sparse learning techniques were implemented on selected features to develop cross-validated models and generate individual-level predictions of antidepressant responses. Response-predictive biomarkers were identified as feature weights of FC-based individual deviations.

We took sertraline as a representative of antidepressants for its widespread usage and abundant pathological studies. First, with an autoencoder-based reconstruction model, we developed a healthy norm of rsEEG connectivity which quantitatively measures the individual deviations of MDD patients in the psychopathological dimensions. Second, to tailor the individual deviations to antidepressant responses, we employed a predictive modeling framework that captures treatment response-correlated individual deviations under supervision, taking advantage of machine learning techniques as they have shown promising capability in precision psychiatry (35). Predictive modeling with the quantified FC deviations was implemented to achieve individual-level prediction of antidepressant responses. Afterward, we confirmed the unique advantage of the individual FC deviation-driven signatures by comparing the results with the predictions yielded by commonly-used clinical measures or raw EEG FCs. We also compared the performance derived from our autoencoder-based individual deviations with other normative modeling strategies, including principal component analysis (PCA)-based and regression-based (36) individual deviations. Moreover, to investigate whether sertraline and placebo treatment responses involve distinct neural circuits, we compared the importance of individual deviations in rsEEG connectivity across treatment arms and brain networks. Finally, we interpreted the autoencoder-based individual deviations using commonly-used clinical measures. Our work aimed to generate translatable antidepressant response prediction models using a novel interpretable normative modeling framework, thus achieving applicable treatment outcome prediction and realizing precision psychiatry for MDD patients.

## Subjects and Methods

### Participants

Overall, we included healthy controls from three independent cohorts and MDD patients from one clinical cohort with antidepressant treatment response data. We integrated three cohorts of healthy controls to ensure our healthy norms represent inter-cohort variability and improve the generalizability of our findings. We focused antidepressant response predictive modeling on one cohort because of the availability of treatment outcome data. A brief summary of subjects included in our study can be found in Supplementary Table 1.

#### Healthy controls

The first cohort consists of 39 healthy subjects from the Establishing Moderators and Biosignatures of Antidepressant Response for Clinical Care (EMBARC) study (37). Subjects were recruited at four study sites (University of Texas Southwestern Medical Center, Massachusetts General Hospital, Columbia University and University of Michigan) with ages between 18 and 65. 8 of these 39 subjects were under 25 (needed to match the demographics with other cohorts). Healthy subjects are defined as both psychiatrically and medically healthy individuals.

The second cohort consists of 75 healthy subjects from the Depression-EEG cohort (38), which was published with an EEG-based analysis of depression and anxiety. These recruited subjects have ages between 18 and 25 with no history of head trauma/seizures and no current use of psychoactive medication. Additionally, healthy subjects are defined by three aspects: (1) consistent low Beck Depression Inventory score (39, 40) (<7) between mass survey and preliminary assessment (2) no self-reported history of MDD, and (3) no self-reported symptoms indicating the possibility of an Axis 1 disorder (41).

The third cohort consists of 153 young healthy subjects (25.1±3.1 years, range 20–35 years, 45 female) from the Leipzig Study for Mind-Body-Emotion Interactions (LEMON) cohort (42, 43) (http://fcon_1000.projects.nitrc.org/indi/retro/MPI_LEMON.html). Excluding subjects without available rsEEG resulted in 138 valid subjects, among which 71 were under age 25. Eligibility for recruitment was determined by two rounds of verification, including a prescreen of prospective participants via telephone and a second individual screening by clinicians. Exclusion criteria included: (1) hypertension diagnosis without intake of antihypertensive medication (2) current and/or previous heart attack or congenital heart defect (3) history (within 10 years) of psychiatric disorders (psychosis, attempted suicide, post-traumatic stress disorder) that required inpatient treatment for longer than 2 weeks (4) history of neurological disorders (multiple sclerosis, stroke, epilepsy, brain tumor, meningoencephalitis, severe concussion) (5) history of malignant diseases (6) consumption of one of the following medications (centrally active medication, beta- and alpha-blocker, cortisol, any chemotherapeutic or psychopharmacological medication) (7) positive drug anamnesis (extensive alcohol, MDMA, amphetamines, cocaine, opiates, benzodiazepine, cannabis) (8) previous participation in scientific study within the last 10 years (9) previous or current enrollment in psychology studies (42).

#### MDD patients

For our study purpose, we only included the 287 MDD patients in the EMBARC study (37), aged between 18 and 65. 21 patients have missing baseline EEG data 45 patients did not return at the end of treatment, resulting in 221 MDD patients available for our study. These valid 221 patients were randomly assigned to a sertraline treatment arm (n = 102) and a placebo treatment arm (n = 119). Inclusion criteria included: (1) having MDD as a primary diagnosis by the Structured Clinical Interview for DSM-IV Axis I Disorders (41) (2) Quick Inventory of Depressive Symptomatology score ≥ 14 (3) a MDD episode beginning before age 30, either a chronic recurrent episode (duration ≥ 2 years) or recurrent major depressive disorder (at least two lifetime episodes) (4) no antidepressant failure during the current episode. Exclusion criteria included: (1) ongoing pregnancy or breastfeeding (2) no use of contraception (3) lifetime history of psychosis or bipolar disorder (4) substance dependence in the past 6 months or substance abuse in the past 2 months (5) unstable psychiatric or general medical conditions requiring hospitalization (6) study medication contraindication (7) clinically significant laboratory abnormalities (8) history of epilepsy or condition requiring an anticonvulsant (9) electroconvulsive therapy, vagal nerve stimulation, TMS or other somatic treatments in the current episode (10) taking medications (including but not limited to antipsychotics and mood stabilizers) (11) ongoing psychotherapy (12) significant suicide risk (13) failure to respond to any antidepressant at adequate dose and duration in the current episode.

For subjects in either treatment arm, an eight-week course of sertraline or placebo was enforced. The random assignment to treatment arms was stratified by site, depression severity, and chronicity. The dosing of medications began at 50mg and was increased to a maximum of 200mg if patients could tolerate and did not respond to lower dosing. The treatment response was evaluated using HAMD_17_. Subjects lacking endpoint HAMD_17_ were excluded from the study for data quality control.

### EEG Data Acquisition and Preprocessing

#### Data acquisition

For the EMBARC cohort, rsEEG was recorded from each of the four study sites, including Columbia University, University of Texas Southwestern Medical Center, University of Michigan, and Massachusetts General Hospital, with varying EEG amplifier settings (37, 44). Specifically, Columbia University collected rsEEG with 72-channels using a 24-bit BioSemi system (sampling rate: 256 Hz, bandpass: DC-251.3 Hz) and a Lycra stretch electrode cap (Electro-Cap International Inc., Ohio). PPO1 and PPO2 were used as the active reference of electrodes. At McLean Hospital, EEG data were collected with 129 channels using a Geodesic Net system (sampling rate: 250 Hz, bandpass: 0.01–100 Hz). Cz was used as the reference (Electrical Geodesics Inc., Oregon). University of Michigan collected EEG data with 60 channels using a 32-bit NeuroScan Synamp (Compumedics, TX) system (sampling rate: 250 Hz, bandpass: 0.5– 100 Hz) and a Lycra stretch electrode cap, with a nose reference. At the University of Texas Southwestern Medical Center, EEG data were collected with 62 channels using the 32-bit NeuroScan Synamp system (sampling rate: 250 Hz, bandpass: DC-100 Hz) and a Lycra stretch electrode cap, with a nose reference. Finally, we used 54 common channels across these four study sites to perform analyses. All four study sites performed amplifier calibrations. rsEEG was recorded in the format of four 2-min blocks (two blocks for eyes-closed and two blocks for eyes open) in a counterbalanced order. During the eyes-open condition, subjects were instructed to remain still, minimize blinks/ eye movements, and fixate on a centrally presented mark on the screen.

For the Depression-EEG cohort (38, 43), participants were instructed to stay relaxed for five minutes during the EEG recording. Signals were collected using 64 Ag/AgCl EEG electrodes (Synamps system) positioned in the standard 10–20 system montage. Signals were sampled at 500 Hz and electrode impedances were kept below 10 kΩ.

For the LEMON cohort (43), participants were asked to be awake during the EEG recording. They were instructed to have their eyes open and fixate on a low-contrast fixation cross on grey background during the eyes-open recording session. For each subject, 16-min resting state EEG was recorded with a ‘BrainAmp plus ’amplifier EEG using 62-channel (with one channel for eye movement) and active ActiCAP electrodes (Brain Products GmbH, Gilching, Germany) positioned in the international standard 10–20 extended localization system. FCz was used as the potential reference of electrodes and the ground was located at the sternum. Electrode impedance was kept below 5 KΩ. Signals were sampled at 2500 Hz.

#### EEG preprocessing

The recorded rsEEG data were cleaned offline with a fully automated artifact rejection pipeline utilized in a previous EEG-based antidepressant response prediction study (44), which aimed to minimize the biases due to subjective manual rejection of artifacts. The entire procedure included the following steps: (1) The resampling of EEG to 250 Hz. (2) The removal of 60 Hz a.c. line noise artifact (45). (3) The removal of non-physiological low frequency in the EEG signals using a 0.01 Hz high-pass filter. (4) The rejection of bad epochs by thresholding the magnitude of each epoch. (5) The rejection of bad channels by thresholding the spatial correlations among channels. (6) The exclusion of subjects with more than 20% bad channels. (7) The estimate of EEG signals from bad channels from the adjacent channels via the spherical spline interpolation (46). (8) An independent component analysis to remove remaining artifacts, including scalp muscle artifact, ocular artifact, and ECG artifact (47). (9) Re-referencing EEG signals to the common average. (10) The filtering of EEG signals to four canonical frequency ranges: theta (4–7 Hz), alpha (8–12 Hz), beta (13–30 Hz), and low gamma (31–50 Hz).

#### Source-Space Connectivity Calculation

With the Brainstorm toolbox (48), we first implemented source localization using the minimum-norm estimation approach (49) to convert the channel-space EEG into the source-space signals of 3,003 vertices. A three-layer (scalp, skull, and cortical surface) boundary element head model was computed with the OpenMEEG plugin (50) based on the FreeSurfer average brain template (51). A total of 3,003 dipoles with unconstrained orientations were generated on the cortical surface. The lead-field matrix relating the dipole activities to the EEG was obtained by projecting the standard electrode positions onto the scalp. For each subject, an imaging kernel that maps from the channel space EEG to the source space current density was then estimated by the minimum norm estimation approach with depth weighting and regularization. Principal component analysis was then employed to reduce the three-dimensional estimated source signal at each vertex to the one-dimensional time series of the principal component.

To capture the brain functional architecture, we extracted connectomic features by calculating power envelope connectivity (PEC)(28) since it has demonstrated strength in mitigating spurious correlations resulting from volume conduction (52). Hilbert transform was first applied to convert source estimates into analytical time series. The analytical time series of each pair of brain signals were then orthogonalized to remove the zero-phase-lag correlation (52). Afterward, the power envelopes were measured by calculating the square of the orthogonalized analytical signals. A logarithmic transform was subsequently conducted to enhance normality. PEC was then calculated as the Pearson’s correlation coefficient between the log-transformed power envelopes of each pair of brain regions. Finally, a Fisher’s r-to-z transformation was performed to enhance normality (43, 52). The regional pairwise PEC features were further extracted based on the Schaefer atlas with 100 brain regions (53). For each pair of regions, connectivity was calculated by averaging PEC values over all possible vertex pairs.

To ensure the individual deviation of each subject was independent of other subjects, the normalization of EEG FC data was conducted on each subject. For each subject, we first performed an inverse hyperbolic tangent transformation on the EEG FC data to better accommodate deviated values of EEG FC. The transformed strengths of 4950 FCs were then z-scored to have a zero mean and unit standard deviation. Notably, this procedure also guaranteed that all the three cohorts had the same FC mean and standard deviation. This normalization strategy endowed the EEG FC of each subject with the independence of other subjects and emphasized the distinguishability between brain regions with high and low FCs.

### Data Harmonization

The utilization of multiple independent cohorts requires data harmonization to eliminate potential site effects due to different data acquisition protocols across studies. The data harmonization was realized with a MATLAB package built on empirical Bayesian methods (ComBat, see references for details) (54–56), which applied a unique transformation on the data from each independent cohorts to project them to a common space. Specifically, the data transformation matrices were first trained using healthy controls and then applied to MDD patients. Therefore, the transformation for MDD patients was independent of the patient population, thus preventing the data leakage issue for the subsequent cross-validation procedure in predictive modeling. Above all, the normalized EEG FC data of healthy controls from the three cohorts (Depression-EEG, EMBARC, LEMON) were combined and used as the input data matrix. The batch labels were assigned based on the cohort origin of each subject, without considering the specific study site within each of the cohorts. Notably, as cohorts had different age ranges, we incorporated age as a covariate in the data harmonization to ensure the eliminated differences were indeed site effects. Eventually, the procedure directly generated the harmonized data of healthy controls, which were subsequently used for normative modeling. For the MDD patients used for treatment response prediction, the same transformation for data harmonization was applied based on their cohort origins without further training or tuning. To demonstrate the importance of data harmonization, we developed prediction models for antidepressant responses using unharmonized EEG data, which showed inferior prediction performance to the model developed with harmonized data (Supplementary Table 2,3). For data harmonization and all subsequent analyses, each EEG condition was processed individually, including all the four frequency bands in either eyes-open or eyes-closed paradigms (totally eight conditions).

### Normative Modeling of Functional Connectivity

Above all, to calibrate demographics across the independent cohorts, thus to improve the reliability and prediction performance of normative modeling, we developed healthy norms of 154 young adults (age 18-25), which consist of 8 subjects from the EMBARC cohort, 75 subjects from the depression-EEG cohort, and 71 subjects from the LEMON cohort. The age threshold of 25 was chosen because the Depression-EEG cohort only had subjects younger than 22, and the LEMON cohort only recorded subjects’ age with a resolution of 5 years. Therefore, including healthy controls with age between 18 and 25 was the best way to match demographics of the three independent cohorts. Healthy norms were derived using three strategies as discussed below.

#### Principal Component Analysis (PCA)-based Modeling

PCA is one of the simplest approach to reconstruct data with lower dimensionality. Here we established the PCA-based reconstruction error to demonstrate that a linear unsupervised reconstruction might be too simple to capture individual deviation information. Specifically, we applied a PCA on the 4950 EEG FCs of healthy subjects and empirically selected the first 100 principal components (explained > 90% variance) to reduce dimensionality. The reconstructed FCs were calculated as the matrix product of kept principal components and transposed eigenvectors. The reconstruction error, which was subsequently used to develop antidepressant response prediction models, was calculated as the difference between original FCs and reconstructed FCs.

#### Regression-based Modeling

Afterward, we calculated individual deviations using an existing normative modeling framework. A recent study developed a comprehensive and user-friendly workflow for realizing normative modeling in precision psychiatry (36). We employed this standardized framework to derive regression-based individual deviations of rsEEG FCs. Briefly, the framework requires users to pre-select a set of covariates and then regress out those covariates based on regression models for FCs. Specifically, we included age and gender as covariates (they were the only two demographic variables shared by all three independent cohorts) and calculated individual deviations in FCs with reference to the age and gender-based FC prediction, which was trained on healthy subjects and applied on MDD patients. The resulting individual deviations were then used to develop antidepressant response prediction models, which unfortunately yielded unsatisfactory prediction performance. Notably, the unsatisfactory performance of regression-based individual deviations in antidepressant response prediction might be due to the lack of common variables that were usable as covariates. It might be also because the regression models were not accurate enough to robustly estimate individual deviations in FCs. But in either case, it demonstrated the approach’s over-reliance on pre-selected covariates and regression performance, calling for an unsupervised normative modeling strategy that is more flexible and accommodating.

#### Autoencoder-based Modeling

The autoencoder has been employed by numerous neuroimaging studies as the normative modeling framework (57–60) for its independence of supervision and high flexibility. The autoencoder we employed for the healthy norm development consists of three layers, including an input layer, a hidden layer, and an output layer. The input and output layers have a dimensionality of 4950, which is the total number of FCs with a 100-ROI brain atlas (53). The hidden layer has a reduced dimensionality to allow the autoencoder to find representative dimensions for the healthy controls (Fig. 1). We empirically selected a dimensionality of 500 for the hidden layer to reduce the dimensionality magnitude while keeping adequate complexity of autoencoders.

The autoencoder training iteratively optimized the parameters for the encoder and the decoder. In practice, we used the MATLAB function “trainAutoencoder” to realize the training process. Specifically, a collection of healthy controls’ EEG FC data was fed into the input layer of the autoencoder. A L2-norm constraint was implemented in the autoencoder training to reduce the collinearity between FCs. Additionally, both encoder and decoder utilize a sigmoid activation function to introduce nonlinearity. The resultant output layer represented the reconstructed EEG FC for each individual subject. Finally, the difference between the original EEG FC and the reconstructed one, termed reconstruction error, was utilized as the quantification of individual deviation from the healthy norm.

As the individual deviations used for sertraline and placebo response prediction were derived from a healthy norm of young adults, we examined whether the prediction performance was dependent on age. Correlation analyses revealed no correlation between sertraline response and age (r = 0.04, p = 0.67, Supplementary Figure 1a), and absolute prediction residuals were also independent of age (EO-alpha: r = -0.09, p = 0.36; EO-gamma: r = -0.09, p = 0.37, Supplementary Figure 1c,d). A further analysis confirmed that the individual deviation dimensions of sertraline response and age were orthogonal (Inner products of age dimension and sertraline response dimension: EO-alpha: -0.007; EO-gamma: -0.028), justifying that the normative modeling based on healthy young adults was capable of predicting sertraline responses of elder MDD patients.

Overall, autoencoder-based individual deviation differed from the previous normative modeling frameworks (31, 32, 61) in three aspects. First, contrary to that previous framework generated predictions for each of the brain regions and then combine them to a data matrix, our framework processed all pairs of ROI-ROI level EEG connectivity together and produces the matrix of individual deviations as a single entity, thus taking interactions between brain regions into account. Second, our framework essentially quantified individual deviations of patients based on a distance mapping of EEG connectivity relative to healthy controls, hence the individual deviation only relies on actual neuroimaging data without dependence on prediction models. Third, autoencoder-based normative modeling is unsupervised, thus not requiring pre-defined covariates and capable of dissecting dimensions not yet discovered by previous studies. Equipped with these features, our normative modeling framework for rsEEG data held the potential to produce a more translatable, objective, generalizable, and robust quantification of individual deviations in psychopathological dimensions.

### Treatment Response prediction

The treatment response prediction was respectively generated for the sertraline and placebo arms, where the treatment outcome was quantified as the pre-minus post-treatment change in HAMD17 score. The general workflow included two steps: feature calculation and predictive modeling. For input features, we compared the performances of raw EEG FCs and normative modeling-quantified individual deviations (Supplementary Tables 4,5). For the predictive modeling, we first applied the connectome-based predictive modeling framework (62) to select connectivity features that were significantly correlated with the outcome. Afterward, as individual deviations in many EEG conditions could predict age (Supplementary Table 6), and age showed modest correlation with placebo response (Supplementary Figure 1), we regressed out age from individual deviation features to ensure that the brain signatures we identified were not confounded by age. Finally, LASSO regression (63, 64)was implemented to further reduce the complexity of final models.

#### Cross-validation for performance evaluation

The performances of prediction models were then evaluated using 10-fold cross-validation. Specifically, the data to be fitted were randomly divided to ten subsets while each subset contained approximately the same subject numbers. One of ten subsets was iteratively held out as the test set while the remaining nine subsets were the training set, such that each subject had a predicted outcome. For LASSO regression, the sparsity parameter *λ* was determined using an inner loop of tenfold cross-validation on the training data. To enhance the stability and reliability of model performance evaluation, the data were randomized ten times and the median of the resulting ten predicted outcomes was used as the final predicted value of each subject. The prediction performance was then quantified as the Pearson’s correlation coefficient and R-squared value between actual and predicted outcomes. The *P* value was calculated against the two-tailed null hypothesis that the Pearson’s correlation coefficient was equal to zero.

#### Connectome-based predictive modeling (CPM)

CPM identifies input features that significantly correlate to the prediction outcome (62), thus reducing the feature dimensionality (65–68). Specifically, we correlated the HAMD_17_ score change with each of the input features (EEG FCs/individual deviations) across all MDD patients in the training set using Pearson’s correlation. Only the input features with significant correlation (threshold P-value = 0.05) to HAMD_17_ score change were retained for the subsequent sparse predictive modeling. To prevent data leakage issues, the CPM procedure was performed inside each fold of cross-validation.

### Relative Importance of Functional Connectivity

To quantify the importance of each FC, we took the absolute values of FC weights in each of the prediction models as both positive and negative weights reflected significant contributions. The absolute feature weights showed different magnitudes for sertraline and placebo response prediction models (Wilcoxon’s rank-sum test: p = 0.0017), with the distribution not following a normal distribution (Kolmogorov-Smirnov test. Sertraline: p = 1.49x 10^-80^; Placebo: p = 7.40x 10^-79^). Thus, we sorted FCs according to their absolute feature weights in each prediction model and used their order indices as the quantification of FC importance to perform non-parametric statistical tests. To remove FCs with very small feature weights, we only incorporated the top 400 FCs in each prediction model, which were approximately the top 50% FCs in each prediction model, corresponding to a z-score threshold of 0.25. Specifically, the FC with the largest/smallest absolute feature weight was assigned with the greatest/lowest importance, which was respectively 400 and 1. Other FCs were assigned with importance by an increment of 1.

## Results

### Individual Deviations of Functional Connectivity Predict Antidepressant Responses

With functional connectivity extracted from pretreatment rsEEG, we built individual deviation-based prediction models of antidepressant responses for each of the eight conditions (frequency bands: theta, alpha, beta, gamma; eye status: open, closed), where individual deviations were derived from an autoencoder-based normative modeling framework (Fig. 1, see Methods for details) and the antidepressant responses were quantified as the pre-minus post-treatment changes in the 17-item Hamilton Depression Rating Scale (69) (HAMD_17_). The prediction analysis with cross-validation found that connectivity deviations in the eyes-open-gamma (EO-gamma) condition significantly predicted treatment responses to sertraline (n = 102, r = 0.43, R-squared = 0.15, p_FDR_ = 4.74 x 10^-5^, Fig. 2a), which significantly outperformed the prediction derived from raw EEG FC (r = 0.17, Fisher’s z = 2.03, p_one-tailed_ = 0.021). The performance yielded by our autoencoder-based individual deviations also outperformed PCA- and regression-based (36) individual deviations for sertraline response prediction using the same predictive modeling framework (PCA-based: r = 0.15, Fisher’s z = 2.17, p_one-tailed_ = 0.015; regression-based: r = 0.20, Fisher’s z = 1.85, p_one-tailed_ = 0.032. Supplementary Figure 2a,b). Individual deviations of connectivity in the EO-alpha condition were also significantly predictive of the sertraline responses (r = 0.33, p_FDR_ = 0.0076, Supplementary Figure 3). However, individual deviations in other EEG conditions were not predictive of sertraline responses (Supplementary Table 7). Remarkably, the best performing sertraline prediction model (EO-gamma) did not predict placebo response (r = 0.03, p = 0.74. Fig. 2b), suggesting the model’s specificity to sertraline response.

**Fig. 2.**
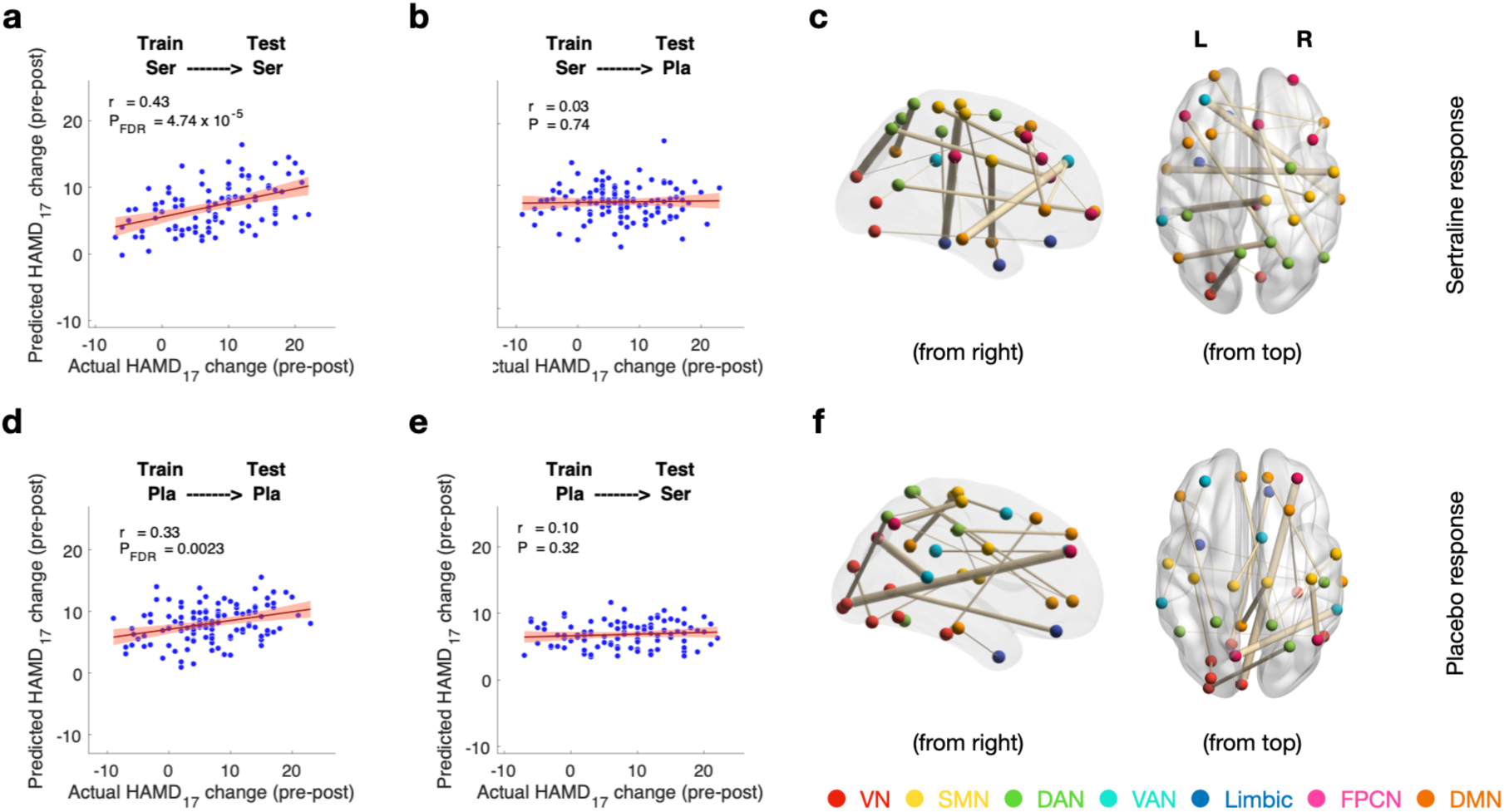
Antidepressant response prediction **a** Performance of the sertraline response model. The model was trained and 10x ten-fold cross-validated on the subjects in the sertraline arm (n = 102). **b** Sertraline response model failed to predict placebo response, demonstrating the model’s specificity for sertraline response prediction. **c** Top 20 significant functional connections for sertraline response prediction **d** Performance of the placebo response model. The model was trained and 10x ten-fold cross-validated on the subjects in the placebo arm (n = 119). **e** Placebo response model failed to predict sertraline response, demonstrating the model’s specificity for placebo response prediction. **f** Top 20 significant functional connections for placebo response prediction. Here, r-values indicate Pearson’s correlation coefficient between actual and predicted antidepressant responses. P-values are calculated based on the two-sided test against the alternative hypothesis of r ≠0. Error bars show standard deviation. VN: visual network; SMN: somatomotor network; DAN: dorsal attention network; VAN: ventral attention network; FPCN: frontoparietal control network; DMN: default-mode network.

For the placebo arm (n = 119), connectivity deviations showed significant predictability of treatment responses in eyes-close-beta (EC-beta) condition (r = 0.33, p_FDR_ = 0.0019, Fig. 2d), which outperformed the raw EEG-FC based model (r = -0.03, Fisher’s z = 2.84, p = 0.002) and the prediction yielded by PCA- and regression-based (36) individual deviations (PCA-based: r = 0.15, Fisher’s z = 1.43, p_one-tailed_ = 0.076; regression-based: r = 0.07, Fisher’s z = 2.07, p_one-tailed_ = 0.019. Supplementary Figure 2c,d). Individual deviations of connectivity in the EC-theta band were also significantly predictive of the placebo response (r = 0.26, p_FDR_ = 0.015) but not in other EEG conditions (Supplementary Table 8). The best placebo-predicting model (EC-beta) could not predict sertraline response (r = 0.10, p = 0.32, Fig. 2e), suggesting the model’s specificity to placebo response.

Subsequently, we examined which FCs were essential to antidepressant response predictions. The top significant connections were identified based on the feature weights derived from the prediction models for sertraline and placebo responses. For sertraline response (as elucidated by EO-gamma condition), the connections between precuneus and superior occipital gyrus, postcentral gyrus and inferior temporal gyrus, precuneus and angular gyrus were the most important connections (Fig. 2c). For placebo response (as elucidated by EC-beta condition), the connections between middle frontal gyrus and calcarine sulcus, orbitofrontal cortex and calcarine sulcus, superior temporal gyrus and precuneus were the most important connections (Fig. 2f). Additionally, we also quantified the importance of network-level connections by averaging the feature weights of non-zero FCs between a particular pair of brain networks. Interestingly, we found a significant and large-scale of involvement of visual and somatomotor networks for placebo response, whereas the involvement of these two networks were sparse and less intense for sertraline response (Fig. 3a,b), implying distinct neural circuits involved in sertraline and placebo responses.

**Fig. 3.**
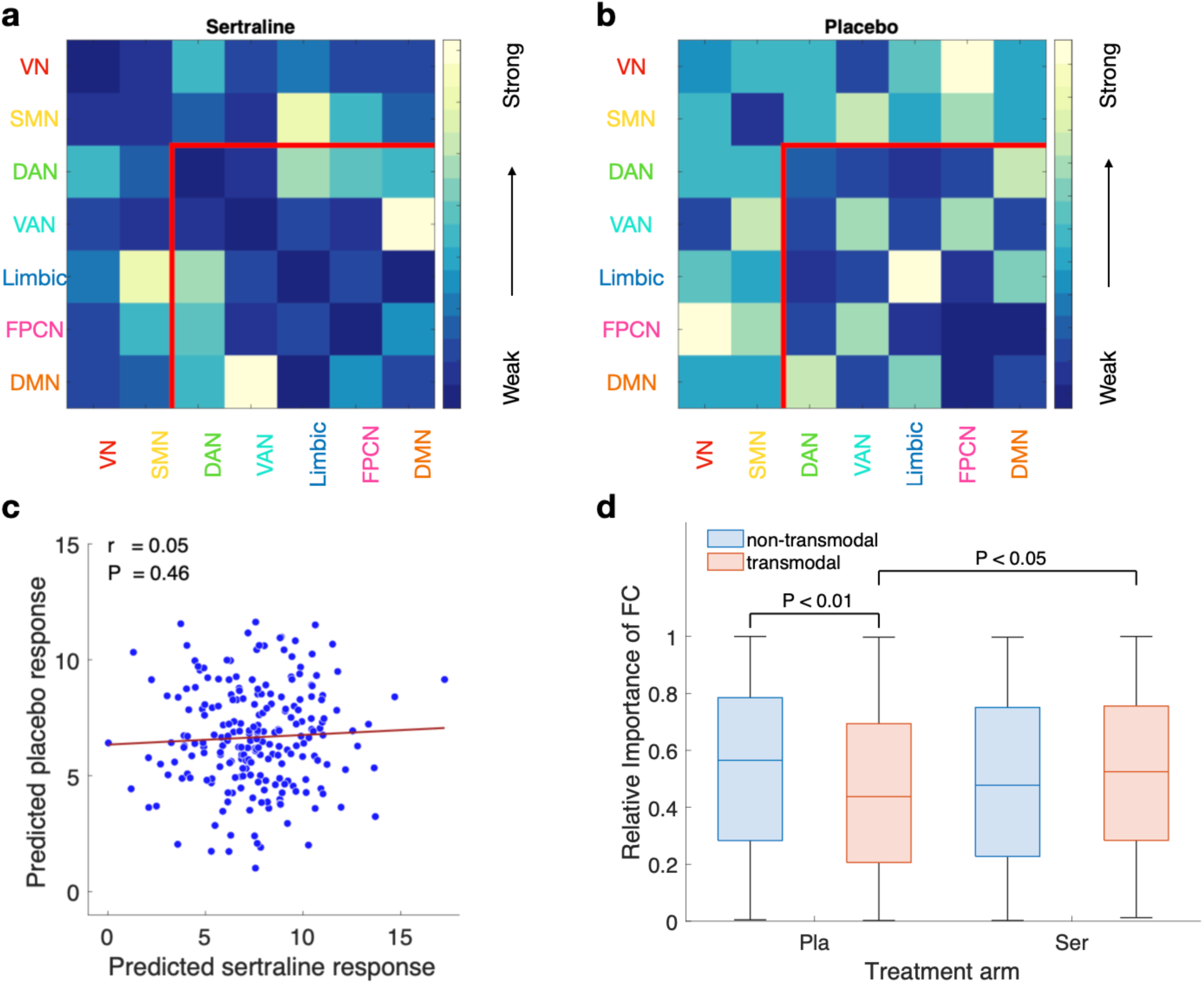
Different pathways underlying sertraline and placebo responses **a-b** Importance of network-level connections for sertraline and placebo responses. Network-level connections were quantified as the mean of non-zero FCs between a particular pair of brain networks. FCs were classified into two groups: the transmodal FCs across transmodal regions (e.g., DMN and FPCN), and the non-transmodal FCs across the unimodal regions (VN and SMN) and between the unimodal regions and transmodal regions. The regions inside the red lines at the bottom-right show the transmodal FCs. The regions outside the red lines show the non-transmodal FCs. **c** Predicted sertraline and placebo responses showed no significant correlation, suggesting the independence between predicted sertraline and placebo responses. The predicted antidepressant responses were acquired for all subjects in the EMMARC cohort (n = 221). **d** Sertraline and placebo responses showed different characteristics in FC weights. The feature weights of transmodal FCs were significantly different between sertraline and placebo responses, whereas no significant difference was identified for the features weights of non-transmodal FCs between sertraline and placebo responses. Moreover, non-transmodal FCs showed significantly higher importance to placebo response prediction than transmodal FCs. These comparisons suggested the relatively higher importance of transmodal FCs to the sertraline response and relatively higher importance of non-transmodal FCs to the placebo response, suggesting different levels of brain networks involved in sertraline and placebo responses. To directly compare the feature weights between two different models, absolute feature weights were ordered for each model, such that the order reflected the relative importance for each of the antidepressant responses. The differences in importance across FC groups were then identified by Wilcoxon’s rank-sum tests.

### Sertraline and Placebo Responses Mediated by Different Hierarchies of Brain Networks

As both sertraline and placebo response prediction models showed specificity to treatment arms, one intriguing question was whether sertraline and placebo responses were achieved via different neurobiological pathways. First, we confirmed the independence between sertraline and placebo responses by performing a correlational analysis on predicted sertraline and placebo responses. Specifically, we acquired predicted antidepressant responses of both sertraline and placebo treatment for all MDD patients — notably, although we could not predict placebo/sertraline response using sertraline/placebo response predictive models, we could predict hypothetical responses should the patients be assigned to the other arm. Afterward, we correlated the predicted sertraline and placebo responses, which yielded no significant correlation (r = 0.05, p = 0.46, Fig. 3c). Additionally, we found that the treatment-predictive phenotype derived from EEG connectivity deviations for sertraline and placebo responses were orthogonal (inner product = -5.23 x 10^-4^). Together, these results suggested that sertraline and placebo responses were independent and might be via distinct pathways.

Next, we investigated the FC signature that predicted sertraline and placebo responses. Inspired by the theory of so-called brain gradients (70, 71), which suggested a hierarchy in brain networks, we classified all FCs into two groups: the transmodal FCs across transmodal regions (e.g., default mode network and frontoparietal networks), and the non-transmodal FCs across the unimodal regions (visual and somatomotor networks) and between the unimodal and transmodal regions. Wilcoxon’s rank-sum tests were performed to identify significant differences of FC importance across treatment arms and the two FC groups (non-transmodal vs. transmodal) (Fig. 3d). As results, non-transmodal FCs showed significantly higher importance than transmodal FCs for predicting placebo response (p = 0.009), and transmodal FCs showed significantly higher importance for predicting sertraline response than placebo response (p = 0.035). These results indicated that placebo response relied heavier on non-transmodal FCs, whereas sertraline response might rely heavier on transmodal FCs, suggesting that placebo response was via more superficial pathways, echoing with the findings that placebo response tends to have earlier onset yet is more prone to relapse and recurrence (72–74).

### Treatment-Predictive Phenotypes Provide Supplementary Information to Commonly-Used Clinical Measures

Neuroimaging-based prediction models may not be practically useful if lower-cost measures (e.g., demographic variables, clinical scores, or historical factors) can yield comparable predictability of antidepressant responses. Therefore, we developed clinical measure-based prediction models for antidepressant responses to confirm the advantages of neuroimaging-based prediction models. We used totally 134 item-level scores in State-Trait Anxiety Inventory (75), Childhood Trauma Questionnaire (76, 77), Quick Inventory of Depressive Symptomatology (78), Mood and Anxiety Symptom Questionnaire (79), and age and education years. These item-level scores yielded no to modest predictability of treatment responses (sertraline response: r = 0.22, p = 0.018; placebo response: r = 0.03, p = 0.75), confirming the advantage of rsEEG-based treatment-predictive phenotypes in indicating antidepressant responses over lower-cost measures (sertraline response: Fisher’s z = 1.84, p = 0.033; placebo response: Fisher’s z = 2.38, p = 0.009).

### Normative Modeling Distinguishes Subclinical and Diagnostic Variabilities

Lastly, as autoencoders reduce feature dimensionality in an unsupervised way, we tested whether indeed the reconstructed rsEEG FC captured the variability in healthy controls and the reconstruction error reflected deviations in psychopathological dimensions. To this end, we correlated 9 clinical measures (five sub-scales of Childhood Trauma Questionnaire (76, 77): emotional abuse, emotional neglect, physical abuse, physical neglect, sexual abuse; total evaluation score of Quick Inventory of Depressive Symptomatology (QIDS) (78); tripartite scales of Mood and Anxiety Symptom Questionnaire (79, 80): general distress, anhedonic depression, anxious arousal) with either reconstructed rsEEG FC or individual deviations of each FC (as elucidated by EO-gamma condition). We compared whether each clinical measure had stronger correlation with reconstructed FCs or individual deviations using paired Wilcoxon’s rank-sum tests, with the absolute correlation coefficients between a particular clinical measure and each FC served as data points (Table 1). To establish the ground truth of whether a clinical measure reflected the variability shared across both healthy controls and patients or the variability specific to patients, we categorized the clinical measures as subclinical or diagnostic depending on whether they significantly distinguish patients from healthy controls (Table 1). Although all clinical measures showed significant distinction between healthy subjects and MDD patients at the significance level of 0.05, two of them (history of physical abuse and sexual abuse) showed a significance level much higher than others (p > 0.0001), thus were categorized as subclinical measures. The other five clinical measures were categorized as diagnostic measures. Interestingly, both subclinical measures showed stronger correlation with reconstructed FCs than individual deviations. Meanwhile, 5 of the 7 diagnostic measures (except the history of emotional abuse and score of anxious arousal) showed stronger correlation with individual deviations than reconstructed FCs. This result suggested that the reconstruction error derived from the autoencoder model indeed distinguished subclinical and diagnostic variabilities, thus verifying the hypothesis that the reconstruction error from normative modeling captures individual deviations in psychopathological dimensions.

**Table 1.** Individual deviation distinguishes subclinical and diagnostic variabilities

|  |  | Correlation between clinical measures and FC feature/diagnosis |  |  |  |
| --- | --- | --- | --- | --- | --- |
|  |  | Absolute correlation difference between reconstructed FC and individual deviation |  | Measure difference between healthy subjects and MDD patients |  |
| Measure |  | z-value | p-value | z-value | p-value |
| CTQ | Emotional abuse | **-6.74** | **1.58 x 10<sup>-11</sup>** | 7.49 | 6.66 x 10<sup>-14</sup> |
|  | Emotional neglect | 6.44 | 1.19 x 10<sup>-10</sup> | 6.94 | 3.83 x 10<sup>-12</sup> |
|  | Physical abuse | **-5.65** | **1.65 x 10<sup>-8</sup>** | **2.68** | **0.0075** |
|  | Physical neglect | 3.26 | 0.0011 | 6.17 | 6.67 x 10<sup>-10</sup> |
|  | Sexual abuse | **-12.99** | **1.49 x 10<sup>-38</sup>** | **3.68** | **0.0002** |
| QIDS | Total score | 12.15 | 5.74 x 10<sup>-34</sup> | 10.13 | 3.94 x 10<sup>-24</sup> |
| MASQ | General distress | 10.95 | 6.56 x 10<sup>-28</sup> | 9.87 | 5.51 x 10<sup>-23</sup> |
|  | Anhedonic depression | 18.55 | 8.57 x 10<sup>-77</sup> | 9.53 | 1.63 x 10<sup>-21</sup> |
|  | Anxious arousal | **-8.78** | **1.69 x 10<sup>-18</sup>** | 8.74 | 2.30 x 10<sup>-18</sup> |

## Discussion

In this study, we developed prediction models for antidepressant responses based on individual deviations from normative rsEEG FC. The prediction models yielded promising performance for both sertraline and placebo responses, demonstrating the potential generalizability of this normative modeling-based prediction framework to other treatment responses. The models outperformed the predictions yielded by rsEEG FC and individual deviations derived from other existing normative modeling strategies (36). Based on the prediction models, we identified distinct connectivity signatures for sertraline and placebo responses, which further suggested differences in involved neural circuits between sertraline and placebo., We showed that the individual deviations indeed provide supplementary information to commonly-used clinical MDD measures for depression, anxiety, childhood history questionnaires, thus unveiling additional information to improve treatment outcome prediction. Finally, we interpreted the autoencoder-based individual deviations using commonly-used clinical measures and verified the hypothesis that the individual deviations indeed distinguish the subclinical and diagnostic variabilities among subjects. Together, these achievements deepened our understanding of how brain FC alterations are associated with antidepressant treatment and a new avenue for improving antidepressant treatment outcomes. Lastly and importantly, this framework is highly generalizable and adaptable to the treatment response prediction studies of other antidepressant medications, and even other psychiatric disorders.

Recently, a number of studies have made efforts in identifying neurobiological biomarkers for antidepressant responses using pretreatment EEG connectivity (81–83). In their reports, connectivity in alpha and gamma bands provided essential information to both antidepressant and placebo responses, among them the parietal regions were the most important (83). Our prediction models also revealed alpha and gamma bands as the most significant conditions for sertraline response, whereas the beta band was the most informative condition for placebo response. More specifically, we found the precuneus region, which is a functional core of the default-mode network (84), of particular importance to the sertraline response. For depression, abnormal activities in precuneus were reported to be associated with low self-esteem (85), recurrence after treatment (86), incidence (87), suicide risk (88), and symptom severity (89). Our predictive modeling results provide additional evidence that precuneus is the key region for antidepressant response, strengthening the finding that precuneus is an important indicator for depression dimensions and verifying the perfect alignment of our study with previous neuroscience studies.

As for placebo response, we found the calcarine sulcus is of particular importance, where the primary visual cortex is concentrated (90), aligning with previous studies that placebo treatment may induce alterations in primary visual cortex (91, 92). We also found that the connections within limbic network were highly involved for placebo response. Interestingly, the alterations in cortical and limbic connectivities we identified for placebo response precisely echoed with neuroimaging biomarkers for psychotherapy (93). Together with our findings, it might imply that the placebo treatment is essentially a special case of psychotherapy, where patients perceive the stimuli of clinicians providing medications to them, as well as clinician’s hint that they would get improvement from the medications.

Notably, in our prediction models, it is the deviations in FC from the healthy norm that contribute to the antidepressant response prediction. In principle, raw EEG FC and individual deviation from normative EEG FC reflect different aspects — raw EEG FC describes the coupling between brain regions, whereas individual deviations in EEG FC represents how the coupling of patients is deviated from healthy conditions. Actually, changes in EEG FC may not lead to alterations in individual deviations as long as the changes are not in the psychopathological dimensions. Therefore, individual deviations eliminate the variance irrelevant to psychopathology, conceptualizing psychopathological dimensions of each individual FC. The significant feature weights in our models essentially indicate the individual deviations from normative connectivity of identified connections are important for predicting treatment responses. As a result, the neurobiological biomarkers we identified from normative modeling provide unique and supplementary information to previous treatment response biomarker studies.

Debates have existed for long about how antidepressant medications take effect on MDD patients (3, 4), as well as the essence of placebo effect and response (11). Our prediction models provided novel insights into their response pathways from the aspect of functional connections. As shown in Results, non-transmodal FCs contribute more to the placebo response than transmodal FCs whereas transmodal FCs contribute more to the sertraline response than placebo response. These comparisons suggest that the more profound effects of sertraline may be because of its higher level of modulation (transmodal regions) than placebo (unimodal regions). It may also explain why sertraline response is only modestly higher than placebo response: they both introduce alterations in brain connectivity, just in different aspects; because the actual behaviors, as measured by clinical scales for MDD, are accomplished by the entire neural pathway including connections involving unimodal and transmodal regions, the overall outcomes are similar. Namely, sertraline and placebo might achieve similar outcomes via distinct neural circuits, which aligned with the results from our simulation analysis that sertraline and placebo response may be independent. Taken together, these observations suggested that placebo may possess a distinct mechanism in releasing depression symptoms, not just being a proportion of drug effects (11).

Although our results were preliminarily verified with aforementioned analyses, replication studies with additional independent cohorts are important to confirm our findings of rsEEG connectivity-based individual deviations and predictive modeling for sertraline and placebo responses. Future studies are also needed to validate the generalizability of our proposed framework to other antidepressants, psychotherapy, neurostimulation therapy, as well as these treatment strategies for other psychiatric disorders. Furthermore, as the original protocol of EMBARC study (37) did not collect neuroimaging data at the end of treatment, we were unable to confirm the brain connectivity alterations mediated by the medications. Therefore, future studies are required to confirm the differences in involved neural circuits between sertraline and placebo responses.

In summary, we developed a novel EEG-based framework that combines normative connectivity modeling and predictive modeling to identify biomarkers of individual antidepressant responses, which yielded promising prediction performance and specificity to treatment arms. Specifically, we quantified individual deviations from the established normative EEG connectivity, which provide supplementary information to existing commonly-used clinical measures. Importantly, the constructed individual deviations were highly interpretable as they showed capability of distinguishing subclinical and diagnostic variabilities across healthy controls and MDD patients. We also identified significant functional connections contributing to antidepressant responses, which showed distinct patterns of sertraline and placebo responses, suggesting distinguishable mechanisms between these two treatments. Together, our work expands the knowledge about the response pathways of antidepressant medications, provides a new avenue toward precision medicine and higher treatment outcomes of MDD, and conceptualizes a highly generalizable framework that may be adapted to studies into other types of antidepressant treatments and a broad range of psychiatric disorders.

## Data Availability

All data produced in the present work are contained in the manuscript.

## Acknowledgements

This work was supported by NIH grant nos. R01MH129694, R21MH130956, and Lehigh University FIG (FIGAWD35), CORE, and Accelerator grants. Portions of this research were conducted on Lehigh University’s Research Computing infrastructure partially supported by NSF Award 2019035. G.A.F. was supported by NIH grant nos. K23MH114023 and R01MH125886 and grants from the Brain and Behavior Research Foundation and One Mind – Baszucki Brain Research Fund. A.E. was supported by NIH grant nos. DP1MH116506 and R44MH123373.

The clinical measures used for interpreting autoencoder-based individual deviations include scores from Childhood Trauma Questionnaire (CTQ), Quick Inventory of Depressive Symptomatology (QIDS) and Mood and Anxiety Symptom Questionnaire (MASQ). For absolute correlation difference between reconstructed FC and individual deviation, z-values are calculated by paired Wilcoxon’s rank-sum test for the absolute correlation coefficients between each clinical measure and either reconstructed FCs or individual deviations in FCs. Negative values indicate stronger correlations with reconstructed FCs than individual deviations, whereas positive values indicate stronger correlations with individual deviations than reconstructed FCs. Positive values hypothetically correspond to the variability in psychopathological dimensions (because individual deviations are supposed to capture it).

For measure difference between healthy subjects and MDD patients, z-values are calculated by unpaired Wilcoxon’s rank-sum test for the measure difference between diagnosis groups. We defined clinical measures p >0.0001 as subclinical, while measures with p-values smaller than 0.0001 as diagnostic. Both subclinical measures, showed stronger correlations with reconstructed FCs than individual deviations, while five of the seven diagnostic measures except (emotional abuse history and anxious arousal) showed stronger correlations with individual deviations than reconstructed FCs, suggesting the healthy norm derived from the autoencoder model indeed distinguished subclinical and diagnostic variabilities, which in turn validated the hypothesis that the reconstruction error from normative modeling captures individual deviations in psychopathological dimensions. P-values are uncorrected.

