## Supplementary Material for "Individual Deviations from Normative Electroencephalographic Connectivity Predict Antidepressant Response"

### Supplementary Information

**Supplementary Figure 1** Relationships of age with actual antidepressant responses and prediction residuals

**Supplementary Figure 2** Antidepressant response prediction with PCA- and regression-based individual deviations

**Supplementary Figure 3** Sertraline response prediction in EO-alpha condition

**Supplementary Table 1** Baseline demographic characteristics of subjects included in this study

**Supplementary Table 2** Sertraline response prediction derived with unharmonized data

**Supplementary Table 3** Placebo response prediction derived with unharmonized data

**Supplementary Table 4** Sertraline response prediction with raw EEG FC features

**Supplementary Table 5** Sertraline response prediction with raw EEG FC features

**Supplementary Table 6** Age prediction with individual deviation features

**Supplementary Table 7** Sertraline response prediction with original individual deviation features

**Supplementary Table 8** Placebo response prediction with original individual deviation features

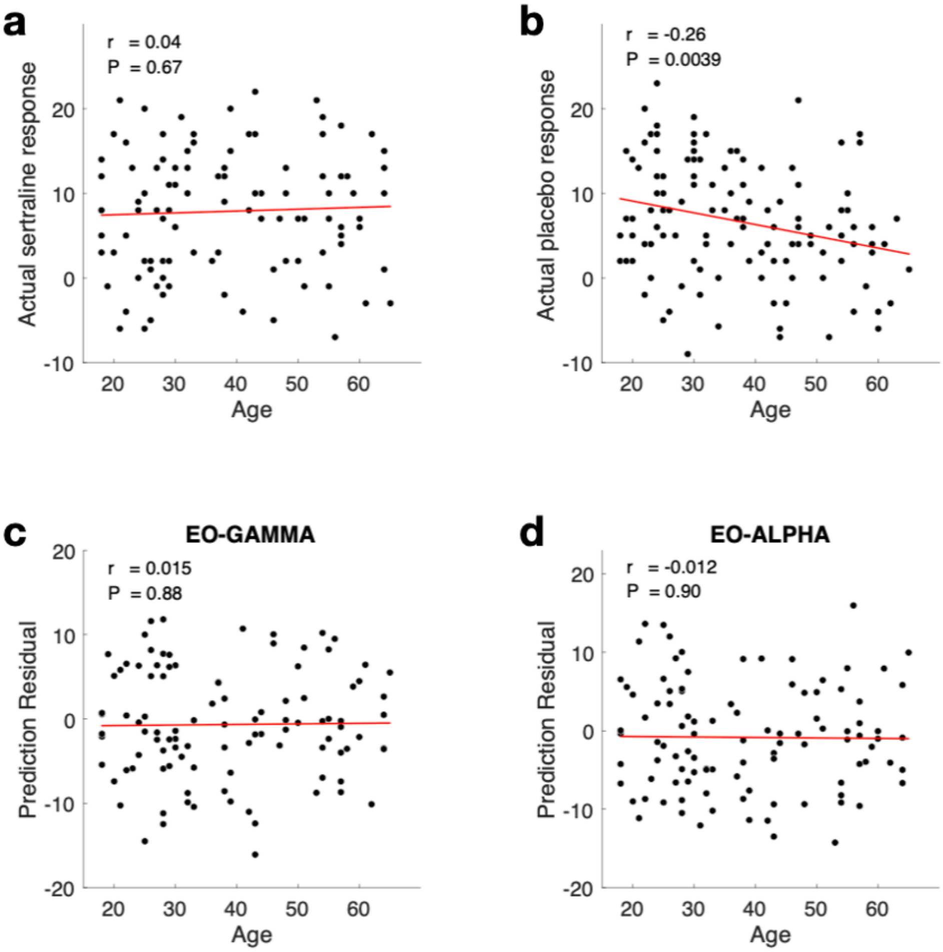

**Supplementary Figure 1** Relationships of age with actual antidepressant responses and prediction residuals. **a** Actual treatment responses in the sertraline arm showed no correlation with age. **b** Actual treatment responses in the placebo arm showed a significant negative correlation with age. **c-d** Prediction residuals of the sertraline response models in EO-gamma and EO-alpha bands showed no correlation with age, confirming the generalizability of the healthy norms to a broad range of ages.

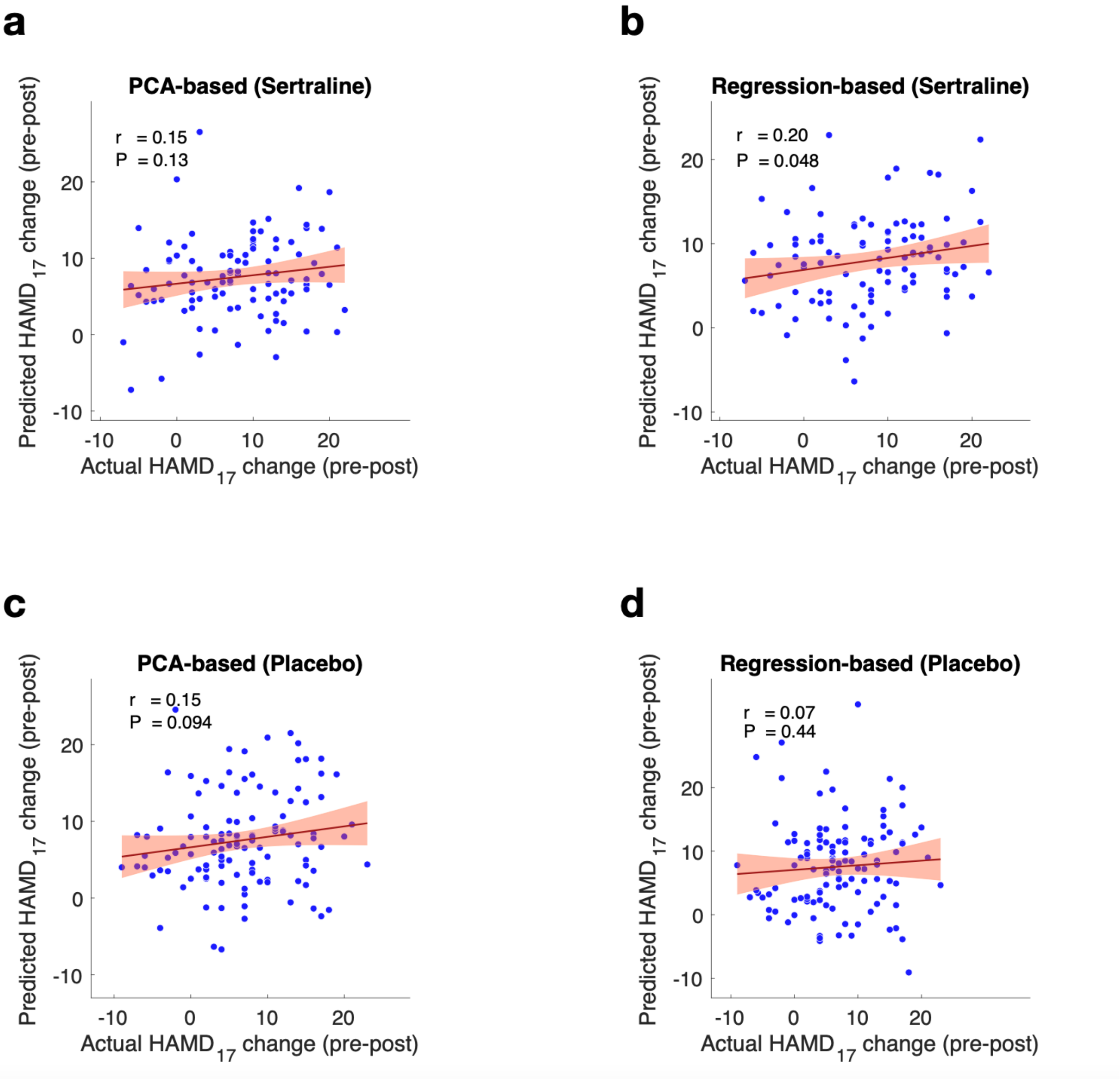

**Supplementary Figure 2** Antidepressant response prediction with PCA- and regression-based individual deviations **a-b** Performance of the sertraline response models (n = 102). **c-d** Performance of the placebo response models (n = 119). P-values are calculated based on the two-sided test against the alternative hypothesis of $r \neq 0$. Error bars show standard deviation.

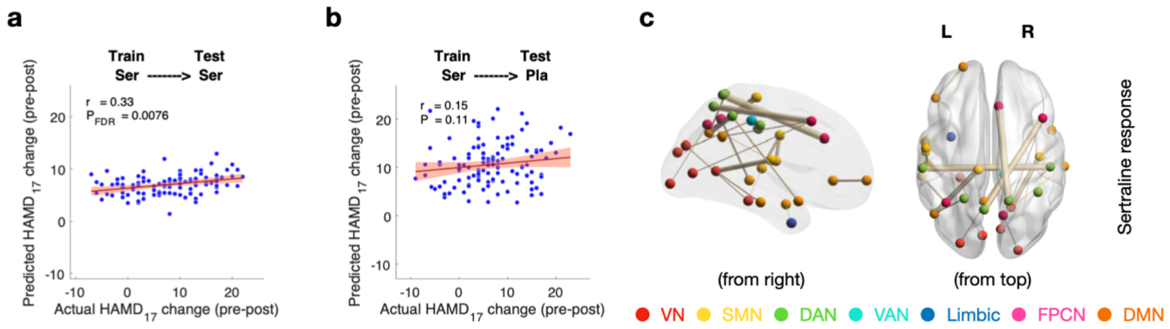

**Supplementary Figure 3** Sertraline response prediction in EO-alpha condition **a** Performance of the sertraline response model. The model was trained and 10x ten-fold cross-validated on the subjects in the sertraline arm (n = 102). **b** Sertraline response model showed insignificant predictability to placebo response, demonstrating the model's specificity for sertraline response prediction. **c** Top 20 significant functional connections for sertraline response prediction in EO-alpha condition. P-values are calculated based on the two-sided test against the alternative hypothesis of $r \neq 0$. Error bars show standard deviation. VN: visual network; SMN: somatomotor network; DAN: dorsal attention network; VAN: ventral attention network; FPCN: frontoparietal control network; DMN: default-mode network.

**Supplementary Table 1** Baseline demographic characteristics of subjects included in this study.

| | Healthy Controls | | | | | MDD patients (EMBARC) | | | |
|---|---|---|---|---|---|---|---|---|---|
| | EMBARC | Depression-EEG | LEMON | | | Sertraline | Placebo | | |
| Categorical Values | (N = 8) | (N = 75) | (N = 71) | $\chi^2$ value | p value | (N = 102) | (N = 119) | $\chi^2$ value | p value |
| Males, No. (%) | 4 (50.0) | 35 (46.7) | 41 (57.8) | 1.81 | 0.40 | 32 (31.4) | 46 (38.7) | 1.28 | 0.26 |
| Continuous variables, mean (SD) | | | | F value | p value | | | t value | p value |
| Age, yr | 21.6 (2.8) | 19.0 (1.2) | 22.5 (NA)* | 0.17 | 1.00 | 38.6 (14.1) | 37.5 (13.0) | 0.61 | 0.54 |
| | * The LEMON cohort only recorded age at the resolution of 5. | | | | | | | | |

**Supplementary Table 2** Sertraline response prediction derived with unharmonized data. LASSO-based sertraline response prediction models were trained and cross-validated in all eight EEG conditions. Normative EEG connectivity was derived from unharmonized data. EEG FCs used for quantifying individual deviations were also unharmonized. Inner loop cross-validation was employed to determine the optimal sparsity parameter for LASSO regression. Possible sparsity parameter values included: 0.01, 0.02, 0.05, 0.1, 0.2, 0.5, 1. The optimal sparsity parameter was determined as the one producing the highest correlation coefficient (r-value). The EEG conditions that generated best performing sertraline prediction models for harmonized data (EO-alpha and EO-gamma) also generated significant models for unharmonized data, but with performance no higher than harmonized data, suggesting the benefits endowed by data harmonization. P-values are corrected using the FDR method.

| | | R-squared | r | $P_{uncorrected}$ | $P_{corrected}$ |
|---|---|---|---|---|---|
| EO | THETA | < 0 | 0.1413 | 0.1567 | 0.2089 |
| | ALPHA | 0.0756 | 0.3296 | $7.17 \times 10^{-4}$ | 0.0029 |
| | BETA | < 0 | 0.0717 | 0.4738 | 0.4877 |
| | GAMMA | 0.1273 | 0.4042 | $2.52 \times 10^{-5}$ | 0.0002 |
| EC | THETA | < 0 | 0.0695 | 0.4877 | 0.4877 |
| | ALPHA | 0.0070 | 0.1501 | 0.1320 | 0.2089 |
| | BETA | < 0 | -0.1946 | 0.0500 | 0.1000 |
| | GAMMA | < 0 | 0.2157 | 0.0294 | 0.0784 |

**Supplementary Table 3** Placebo response prediction derived with unharmonized data. LASSO-based placebo response prediction models were trained and cross-validated in all eight EEG conditions. Normative EEG connectivity was derived from unharmonized data. EEG FCs used for quantifying individual deviations were also unharmonized. Inner loop cross-validation was employed to determine the optimal sparsity parameter for LASSO regression. Possible sparsity parameter values included: 0.01, 0.02, 0.05, 0.1, 0.2, 0.5, 1. The optimal sparsity parameter was determined as the one producing the highest correlation coefficient (r-value). The EEG conditions that generated the best performing sertraline prediction model for harmonized data (EC-beta) also generated significant models for unharmonized data, as well as the EO-theta and EO-gamma conditions. But their performances were lower than the optimal performance derived from harmonized data, suggesting the benefits endowed by data harmonization. P-values are corrected using the FDR method.

| | | R-squared | r | $P_{uncorrected}$ | $P_{corrected}$ |
|---|---|---|---|---|---|
| EO | THETA | 0.0547 | 0.2507 | 0.0060 | 0.0240 |
| | ALPHA | < 0 | -0.0140 | 0.8798 | 0.9988 |
| | BETA | < 0 | 0.0147 | 0.8741 | 0.9988 |
| | GAMMA | 0.0118 | 0.2335 | 0.0106 | 0.0283 |
| EC | THETA | < 0 | -0.0885 | 0.3384 | 0.5414 |
| | ALPHA | < 0 | 0.1425 | 0.1221 | 0.2442 |
| | BETA | 0.0621 | 0.2922 | 0.0013 | 0.0104 |
| | GAMMA | < 0 | -0.0001 | 0.9988 | 0.9988 |

**Supplementary Table 4** Sertraline response prediction with raw EEG FC features. LASSO-based sertraline response prediction models were trained and cross-validated in all eight EEG conditions. Inner loop cross-validation was employed to determine the optimal sparsity parameter for LASSO regression. Possible sparsity parameter values included: 0.01, 0.02, 0.05, 0.1, 0.2, 0.5, 1. The optimal sparsity parameter was determined as the one producing the highest correlation coefficient (r-value). Raw EEG FC-based models only yielded unsatisfying performance for sertraline response prediction which is lower than the predictions derived from individual deviations from normative EEG connectivity. P-values are corrected using the FDR method.

| | | R-squared | r | $P_{uncorrected}$ | $P_{corrected}$ |
|---|---|---|---|---|---|
| EO | THETA | < 0 | -0.0902 | 0.3675 | 0.5880 |
| | ALPHA | < 0 | 0.2873 | 0.0034 | 0.0136 |
| | BETA | < 0 | -0.0464 | 0.6432 | 0.8218 |
| | GAMMA | < 0 | 0.2178 | 0.0279 | 0.0744 |
| EC | THETA | < 0 | 0.0204 | 0.8390 | 0.8390 |
| | ALPHA | < 0 | -0.0361 | 0.7191 | 0.8218 |
| | BETA | < 0 | -0.3208 | 0.0010 | 0.0080 |
| | GAMMA | 0.0040 | 0.1426 | 0.1527 | 0.3054 |

**Supplementary Table 5** Placebo response prediction with raw EEG FC features. LASSO-based placebo response prediction models were trained and cross-validated in all eight EEG conditions. Inner loop cross-validation was employed to determine the optimal sparsity parameter for LASSO regression. Possible sparsity parameter values included: 0.01, 0.02, 0.05, 0.1, 0.2, 0.5, 1. The optimal sparsity parameter was determined as the one producing the highest correlation coefficient (r-value). Raw EEG FC-based models only yielded unsatisfying performance for placebo response prediction which is lower than the predictions derived from individual deviations from normative EEG connectivity. P-values are corrected using the FDR method.

| | | R-squared | r | $P_{uncorrected}$ | $P_{corrected}$ |
|---|---|---|---|---|---|
| EO | THETA | 0.0144 | 0.1667 | 0.0701 | 0.1869 |
| | ALPHA | < 0 | 0.0211 | 0.8199 | 0.8199 |
| | BETA | < 0 | -0.1449 | 0.1159 | 0.2045 |
| | GAMMA | < 0 | 0.1748 | 0.0573 | 0.1869 |
| EC | THETA | < 0 | -0.1875 | 0.0411 | 0.1869 |
| | ALPHA | < 0 | 0.1404 | 0.1278 | 0.2045 |
| | BETA | < 0 | -0.0313 | 0.7356 | 0.8199 |
| | GAMMA | < 0 | 0.0107 | 0.8016 | 0.8199 |

**Supplementary Table 6** Age prediction with individual deviation features. LASSO-based age prediction models were trained and cross-validated in all eight EEG conditions. Inner loop cross-validation was employed to determine the optimal sparsity parameter for LASSO regression. Possible sparsity parameter values included: 0.01, 0.02, 0.05, 0.1, 0.2, 0.5, 1. The optimal sparsity parameter was determined as the one producing the highest correlation coefficient (r-value). Because we aimed to utilize the age dimension to calibrate individual deviation features for antidepressant response predictions, the goodness-of-fit of age prediction model, quantified as R-squared values, was of particular importance. Therefore, we used the R-squared threshold of 0.1 to identify practically useful models, which suggested models in EO-alpha, EC-theta, EC-alpha, and EC-beta conditions. Although EO-beta and EO-gamma also yielded statistically significant age prediction models, the R-squared values of their models were not adequately high to achieve robust age calibration for individual deviation features. P-values are corrected using the FDR method.

| | | R-squared | r | $P_{uncorrected}$ | $P_{corrected}$ |
|---|---|---|---|---|---|
| EO | THETA | < 0 | 0.0230 | 0.7337 | 0.8385 |
| | **ALPHA** | **0.1559** | **0.3997** | **$6.97 \times 10^{-10}$** | **$2.79 \times 10^{-9}$** |
| | BETA | 0.0793 | 0.2961 | $7.53 \times 10^{-6}$ | $1.20 \times 10^{-5}$ |
| | GAMMA | 0.0245 | 0.2914 | $1.07 \times 10^{-5}$ | $1.43 \times 10^{-5}$ |
| EC | **THETA** | **0.1049** | **0.3480** | **$1.09 \times 10^{-7}$** | **$2.18 \times 10^{-7}$** |
| | **ALPHA** | **0.1401** | **0.3770** | **$7.13 \times 10^{-9}$** | **$1.90 \times 10^{-8}$** |
| | **BETA** | **0.1856** | **0.4459** | **$3.39 \times 10^{-12}$** | **$2.71 \times 10^{-11}$** |
| | GAMMA | < 0 | 0.0022 | 0.9737 | 0.9737 |

**Supplementary Table 7** Sertraline response prediction with individual deviation features. LASSO-based sertraline response prediction models were trained and cross-validated in all eight EEG conditions. Inner loop cross-validation was employed to determine the optimal sparsity parameter for LASSO regression. Possible sparsity parameter values included: 0.01, 0.02, 0.05, 0.1, 0.2, 0.5, 1. The optimal sparsity parameter was determined as the one producing the highest correlation coefficient (r-value). The EO-gamma band yielded the best performing model. The EO-alpha band also yielded a significant model for sertraline response prediction. P-values are corrected using the FDR method.

| | | R-squared | r | $P_{uncorrected}$ | $P_{corrected}$ |
|---|---|---|---|---|---|
| EO | THETA | < 0 | 0.1034 | 0.3008 | 0.3438 |
| | ALPHA | 0.0683 | 0.3303 | 0.0019 | 0.0076 |
| | BETA | < 0 | -0.0706 | 0.4810 | 0.4810 |
| | GAMMA | 0.1451 | 0.4316 | $5.92 \times 10^{-6}$ | $4.74 \times 10^{-5}$ |
| EC | THETA | < 0 | 0.1198 | 0.2302 | 0.3069 |
| | ALPHA | < 0 | 0.1217 | 0.2230 | 0.3069 |
| | BETA | < 0 | -0.1928 | 0.0522 | 0.1312 |
| | GAMMA | < 0 | 0.1830 | 0.0656 | 0.1312 |

**Supplementary Table 8** Placebo response prediction with individual deviation features. LASSO-based placebo response prediction models were trained and cross-validated in all eight EEG conditions. Inner loop cross-validation was employed to determine the optimal sparsity parameter for LASSO regression. Possible sparsity parameter values included: 0.01, 0.02, 0.05, 0.1, 0.2, 0.5, 1. The optimal sparsity parameter was determined as the one producing the highest correlation coefficient (r-value). The EO-theta band yielded a significant, yet not satisfactory model. P-values are corrected using the FDR method.

| | | R-squared | r | $P_{uncorrected}$ | $P_{corrected}$ |
|---|---|---|---|---|---|
| EO | THETA | 0.0614 | 0.2637 | 0.0038 | 0.0152 |
| | ALPHA | < 0 | 0.0574 | 0.5356 | 0.5489 |
| | BETA | < 0 | 0.0555 | 0.5489 | 0.5489 |
| | GAMMA | < 0 | 0.1862 | 0.0426 | 0.1136 |
| EC | THETA | < 0 | -0.0619 | 0.5035 | 0.5489 |
| | ALPHA | < 0 | 0.1135 | 0.2192 | 0.4384 |
| | BETA | 0.0060 | 0.3309 | $2.37 \times 10^{-4}$ | 0.0019 |
| | GAMMA | < 0 | -0.0635 | 0.4929 | 0.5489 |
